# Outcomes of a resource-adapted Wilms tumor treatment protocol in Lilongwe, Malawi, 2016-2021: successes and enduring barriers to cure

**DOI:** 10.1101/2022.08.08.22278537

**Authors:** David M. Holmes, Apatsa Matatiyo, Atupele Mpasa, Minke H. W. Huibers, Geoffrey Manda, Tamiwe Tomoka, Maurice Mulenga, Ruth Namazzi, Parth Mehta, Mark Zobeck, Rizine Mzikamanda, Murali Chintagumpala, Carl Allen, Jed G. Nuchtern, Eric Borgstein, Daniel C. Aronson, Nmazuo Ozuah, Bip Nandi, Casey L. McAtee

## Abstract

**Purpose:** Wilms tumor is a common renal cancer of childhood with long-term survival rates exceeding 80% in high-resource countries, yet survival remains below 50% in the low-resource settings of Africa. We assessed outcomes of a resource-adapted treatment protocol at a Malawian hospital to identify actionable factors affecting survival.

**Methods:** We assessed clinical outcomes with a single-center retrospective cohort study of children diagnosed between 2016 and 2021 in Lilongwe, Malawi.

**Findings:** We identified 136 patients with Wilms tumor, most commonly with stage III (25.7%) or IV disease (25.7%). Two-year overall survival (OS) was: Stage I, 78%; Stage II, 27%; Stage III, 62%; Stage IV, 23%, Stage V, 0%. Event-free survival (EFS) was: Stage I, 60%; Stage II, 0%; Stage III, 51%; Stage IV, 13%; Stage V, 0%. After death, treatment abandonment was the most common event comprising EFS, occurring in 26.5% of patients. Among 43% of patients who completed therapy, 2-year OS was 80% and EFS was 69%. Relapse was documented in 9.6% of patients. Radiotherapy was indicated for 40.4% patients, among whom only three received it due to regional unavailability. Factors associated with OS were severe acute malnutrition (Hazard ratio, HR, 1.9), increasing tumor stage (HR, 1.5), and inferior vena cava involvement (HR, 2.7). On multivariable analysis, only tumor stage remained associated with outcome.

**Interpretation:** Implementing a curative resource-adapted treatment protocol in an extremely resourced-constrained environment was feasible in Malawi and resulted in relatively favorable outcomes in low-stage disease, particularly among those who completed therapy. However, factors such as late-stage disease, frequent abandonment, and absent radiotherapy represent ongoing implementation barriers that should be the focus of continued research funding and intervention in Africa.

## Introduction

Wilms tumor (nephroblastoma), an embryonal tumor of the kidney, is among the most common cancers of childhood.^1,2^ In high-income countries, advances in therapy have achieved long-term survival in >80% of children.^3,4^ This success is attributable to early diagnosis, multidisciplinary risk-adapted therapy, and well-resourced supportive care infrastructure.^3,5,6^ Similar outcomes have not been realized in low-and-middle income countries (LMICs) where most pediatric cancer occurs, particularly in sub-Saharan Africa where long-term survival remains dismal.^7–9^ A systematic review of Wilms tumor outcomes in the LMICs of sub-Saharan Africa from 2000-2019 found that overall survival was consistently between 20-50%.^2^

Such disparities are the result of limited capacity to deliver pediatric cancer care within the context of resource-constrained public health systems. Capacity limitations manifest as delayed diagnosis, treatment abandonment, personnel shortages, absent therapeutics, and limited supportive care infrastructure.^1,10^ To overcome these challenges, pediatric cancer centers in LMICs have implemented resource-adapted strategies that contextually adapt protocols to local capacity constraints, such as by lowering chemotherapy intensity to limit blood product requirements.^11–13^

Wilms tumor is treated with chemotherapy, surgery, and radiotherapy, but access to these components remains unavailable throughout much of Africa.^3,5^ The purpose of this study was to determine the outcomes of a pragmatic, resource-adapted Wilms tumor protocol in a cancer center in Malawi, a low-income country with limited medical infrastructure typical of countries throughout sub-Saharan Africa. We also aimed to identify actionable factors influencing mortality to continue to improve the treatment approach to Wilms tumor in similar settings.

## Materials and Methods

### Study design and setting

We conducted a retrospective cohort study of patients aged ≤18 years with Wilms tumor between January 2016 and December 2021 at Kamuzu Central Hospital (KCH) in Lilongwe, Malawi, a referral center serving a population of nine million people spanning Central and Northern Malawi. The Pediatric Oncology Unit contains 30 beds with access to chemotherapy, pathology services, oncology nursing, pediatric hematologist-oncologists, and pediatric surgeons.

### Patient characteristics and diagnosis

Data were extracted from paper charts. Tumor stage and histologic grade were determined according to International Society of Paediatric Oncology (SIOP) guidelines by reviewing histological and surgical reports.^14^ In accordance with SIOP guidelines, patients were identified by characteristic abdominal imaging. These guidelines implement a post-surgical staging system; thus, histological and surgical evaluation are required to differentiate local stages I-III (Supplemental table 1). Patients with Stage IV and V disease were identified with imaging; otherwise, patients without surgical or histopathological data available (e.g., due to presurgical death) were categorized as un-staged. Malnutrition was diagnosed and treated by nutritionists according to World Health Organization guidelines.^15^

### Treatment received

The treatment protocol was built upon a SIOP backbone incorporating eight weeks of pre-operative vincristine (1.5 mg/m^2^/dose) and dactinomycin (0.045 mg/kg/dose) for non-metastatic disease with the addition of doxorubicin (50 mg/m^2^/dose) for metastatic or bilateral disease. Standard chemotherapy dose adjustments were made for children <12 months.^16^ Post-operative chemotherapy regimens were selected according to tumor stage and tumor histology (Supplemental table 2). Surgical complications were defined as any of the following during or up to 30 days post-operatively: death, re-operation, surgical site infection, or wound dehiscence.

Patients with persistent metastatic disease at the time of nephrectomy and patients with Stage V disease were given escalated treatment intensity with cyclophosphamide (2200 mg/m^2^/cycle) and etoposide (500 mg/m^2^/cycle) added to vincristine (2 mg/m^2^/dose) and doxorubicin (30 mg/m^2^/dose) (i.e., Children’s Oncology Group “Regimen M”).^4^ If patients were deemed unlikely to tolerate escalated treatment intensity, the pre-operative regimen was continued. While radiotherapy is not available in Malawi, need for radiation was determined consistent with SIOP guidelines.^17^

The protocol was modified to accommodate resource limitations consistent with SIOP published guidelines for LMICs.^12^ Specifically, pre-operative chemotherapy was extended for up to eight weeks (12 weeks for Stage V) to minimize risk of perioperative morbidity/mortality. Two additional site-specific modifications were made: 1) Patients with Stage I or II disease with low-risk histology were given five cycles of dactinomycin and vincristine-based therapy due to limited relapse salvage options in Malawi, and 2)

Selected patients with Stage III tumors who tolerated pre-operative chemotherapy were chosen to receive Regimen M post-operatively. Finally, upon frequent stockouts of dactinomycin, dactinomycin-containing regimens were substituted during these periods with doxorubicin (50 mg/m^2^/cycle) and cyclophosphamide (1200 mg/m^2^/cycle).

### Statistical analysis

Overall survival was determined by vital status at patients’ last encounter. A treatment-related death was defined as death during therapy unattributable to disease progression. Event-free survival was calculated using the earliest of death, disease progression, relapse, or treatment abandonment. Treatment abandonment was defined as an unplanned hiatus of ≥4 weeks in the scheduled curative-intent therapy plan.^18^ Patients were right-censored at the earlier of their last date of follow-up or March 2022.

Kaplan-Meier survival curves and Cox proportional hazards models were used to identify risk factors associated with survival. Variables assessed were age, sex, use of traditional medicine, acute malnutrition at diagnosis, tumor stage, time-to-surgery, and inferior vena cava involvement. Median follow-up time was calculated using the reverse Kaplan-Meier method.^19^

Analyses were performed in R.^20,21^ Data collection occurred from January-March 2022 with analysis through May 2022 in accordance with Strengthening the Reporting of Observational Studies in Epidemiology (STROBE) guidelines.^22^ The study was approved by institutional review boards in Malawi and the United States.

## Results

### Study cohort

We identified 141 patients with suspected Wilms tumor. Five patients (3.5%) with non-Wilms histology were excluded to create a study cohort of 136 patients (Table 1). Non-Wilms diagnoses included neuroblastoma, renal cell carcinoma, and mesoblastic nephroma. The median age was 3.8 years (interquartile range, IQR, 2.4-5.3). A single patient was HIV positive.

**Table 1:** Characteristics of 136 patients with Wilms tumor within the study cohort.

|  |  |
| --- | --- |
| **Age in years (Median, IQR)** | 3.8 (2.4, 5.3) |
| **Male sex** | 68 (50.0%) |
| **Reported duration of symptoms in weeks (median, IQR)** | 6.0 (4.0, 12.0) |
| **Moderate acute malnutrition at diagnosis** | 20 (14.7%) |
| **Severe acute malnutrition at diagnosis** | 36 (26.5%) |
| **Overall stage** |  |
| Stage I | 30 (22.1%) |
| Stage II | 7 (5.1%) |
| Stage III | 35 (25.7%) |
| Stage IV | 35 (25.7%) |
| Stage V | 5 (3.7%) |
| Un-staged | 24 (17.6%) |
| **Wilms histological subtype<sup>^</sup>** |  |
| Low risk | 8 (11%) |
| Intermediate risk | 56 (77%) |
| High risk | 9 (12%) |

**Legend:** IQR, interquartile range. Data are n (%) unless otherwise specified.
<sup>^</sup> Histologic subtypes are 1) Low risk: 100% necrotic; 2) Intermediate risk: regressive type, mixed type, epithelial type, stromal type, or focal anaplasia; and 3) High risk: blastemal type or diffuse anaplasia. Histological grade data represent 73 patients with post-surgical histological grade data available.

Radiological diagnoses were made by ultrasound in 123 patients (90.4%) and by computed tomography (CT) with/without ultrasound in 63 (46.3%). Chest imaging was documented by X-ray (n=98) and/or CT (n=76) in 126 patients (92.7%). The median reported duration of symptoms prior to presentation was six weeks (IQR 4-12). Moderate acute malnutrition was identified in 20 (14.7%) patients and severe acute malnutrition in 36 (26.5%) (Table 1).

### Tumor characteristics and staging

Most patients had either Stage III (25.7%) or Stage IV (25.7%, Table 1) disease. Staging was unavailable for 24 (17.7%) patients, most commonly due to pre-operative death or treatment abandonment. Bilateral disease occurred in five patients (3.7%). Among those with metastatic disease, 16 (11.8%) had pulmonary metastases, 10 (7.4%) hepatic, and 7 (5.1%) both. Thirteen (9.6%) patients had tumor within the IVC. Post-operative histology reports were available for 74 patients (80% of patients undergoing nephrectomy). Among these, 8 (10.8%) were SIOP low risk tumors, 56 (75.7%) intermediate risk, and 9 (12.2%) high risk.

### Therapy received

Pre-operative chemotherapy was started in 129 (94.9%) patients (Figure 1). Two patients had up-front resection for suspected mesoblastic nephroma, and five patients were discharged on palliative care. The median duration of pre-operative chemotherapy (i.e., time-to-surgery) was seven weeks (IQR, 5-11). Ninety-two patients (68%) underwent nephrectomy. The most common reasons for failure to reach nephrectomy were pre-operative death (50%) and treatment abandonment (39%).

**Figure 1:**
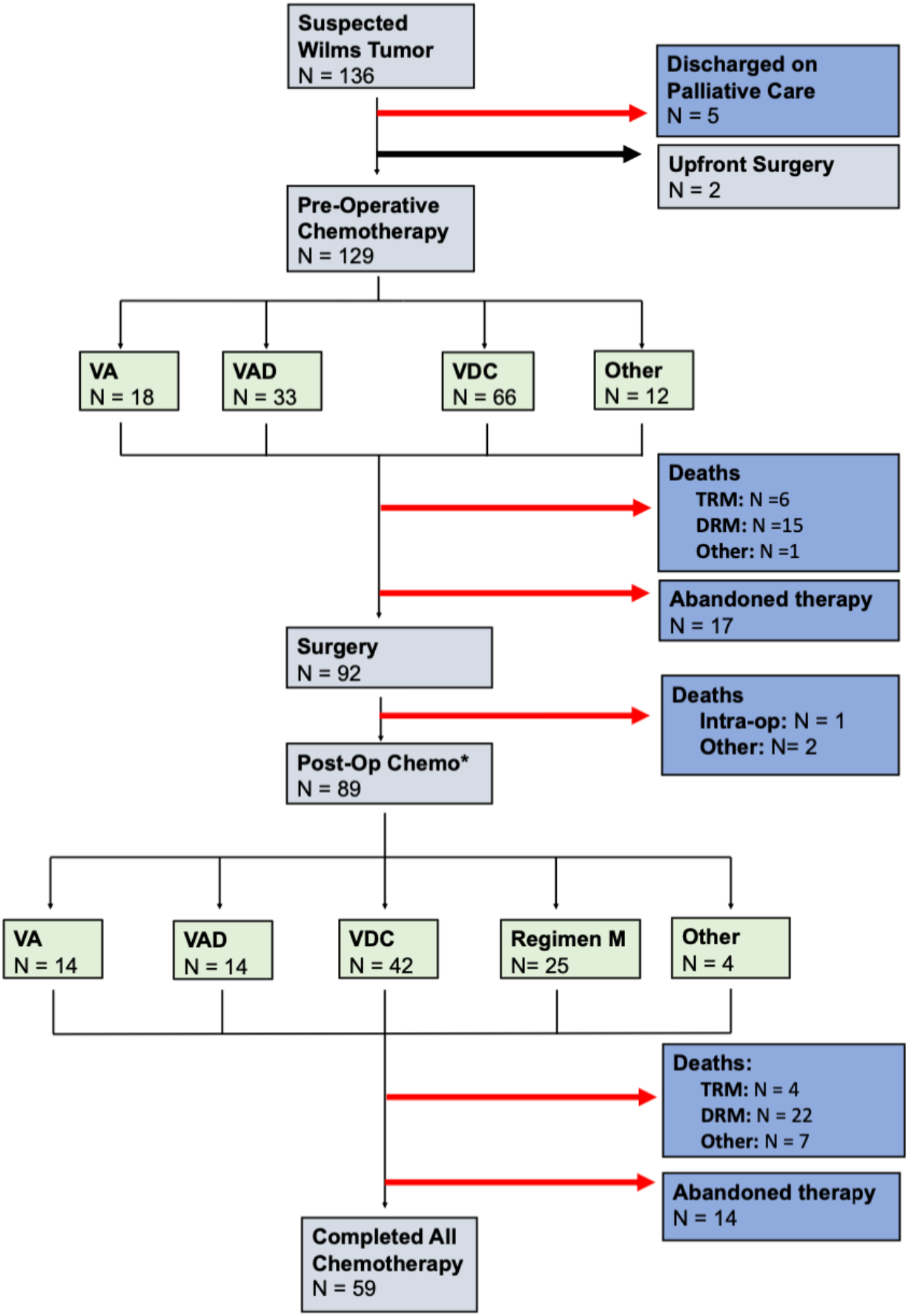
Study profile for Wilms tumor Patients treated at KCH from 2016-2021. T **Legend:** The number of abandoned patients depicted does not include those who abandoned but returned to continue therapy. TRM = treatment related mortality, DRM = disease related mortality, VA = vincristine and dactinomycin, VAD = vincristine, dactinomycin, and doxorubicin, VDC = vincristine, doxorubicin, and cyclophosphamide

Eighty-nine patients (65.4%) received postoperative chemotherapy for a median duration of 15 weeks (IQR, 10-22 weeks; Table 2). In total, fifty-nine patients (43.4%) completed therapy. The most common reasons for failure to complete therapy were death and treatment abandonment. Thirty-six patients (26.5%) abandoned therapy. The median time to abandonment was eight weeks (IQR, 1-17 weeks), with 53% abandoning prior to nephrectomy.

**Table 2.**
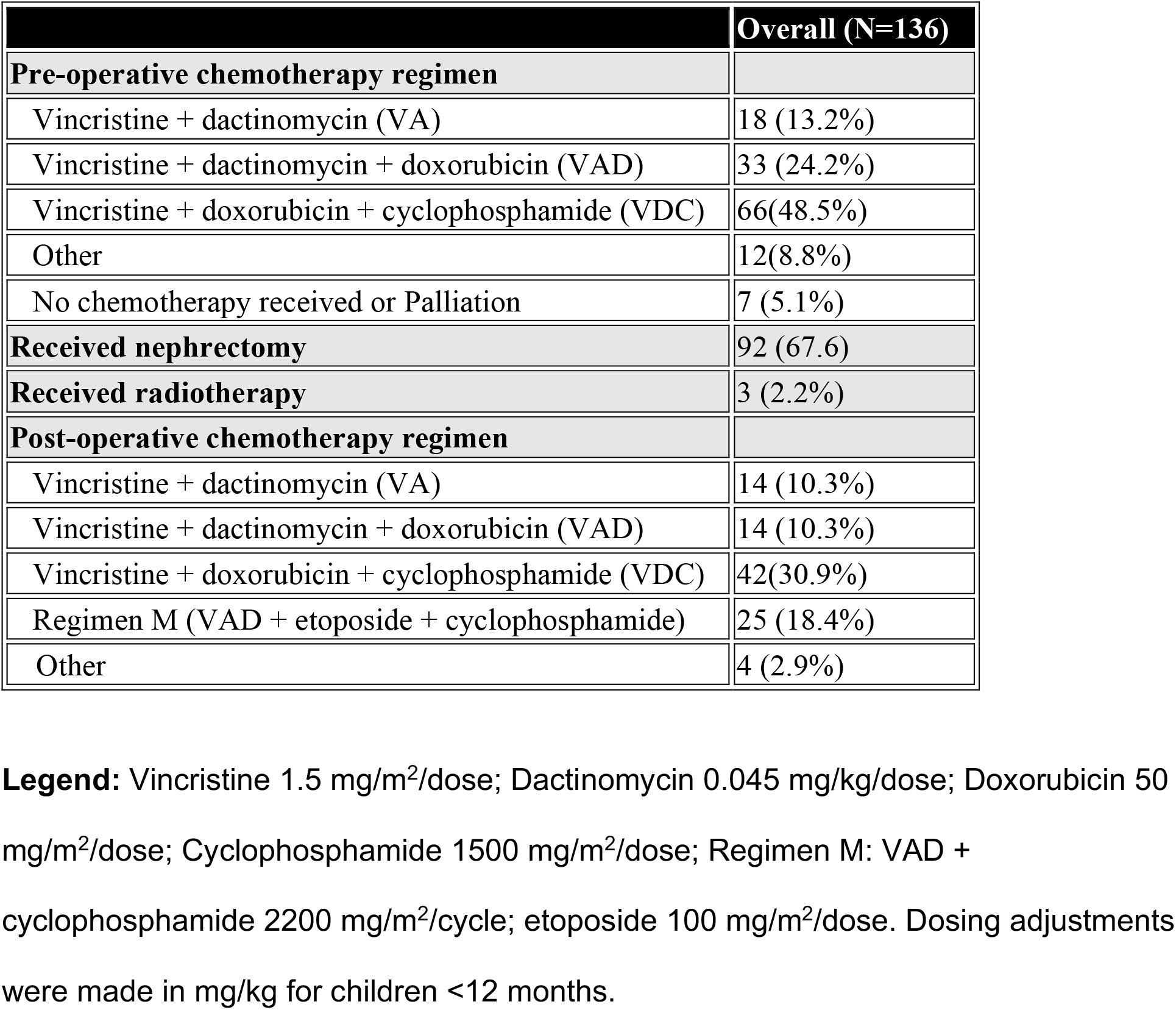
Chemotherapy regimens, surgery, and radiotherapy received.

Radiotherapy was indicated for 55 patients (40.4%). Among these, three received it abroad due to its unavailability in Malawi.

### Surgical outcomes

Operative or postoperative complications occurred in 12 patients (13% of nephrectomies), most often due to infection (Table 3). The 30-day re-operation rate was 4%. There were four cases of post-operative intussusception requiring laparotomy and reduction, and one case of small bowel stricture requiring laparotomy with bowel resection and anastomosis. A single intra-operative death occurred due to suspected hemorrhage versus air embolus. There were no other 30-day perioperative deaths.

**Table 3:**
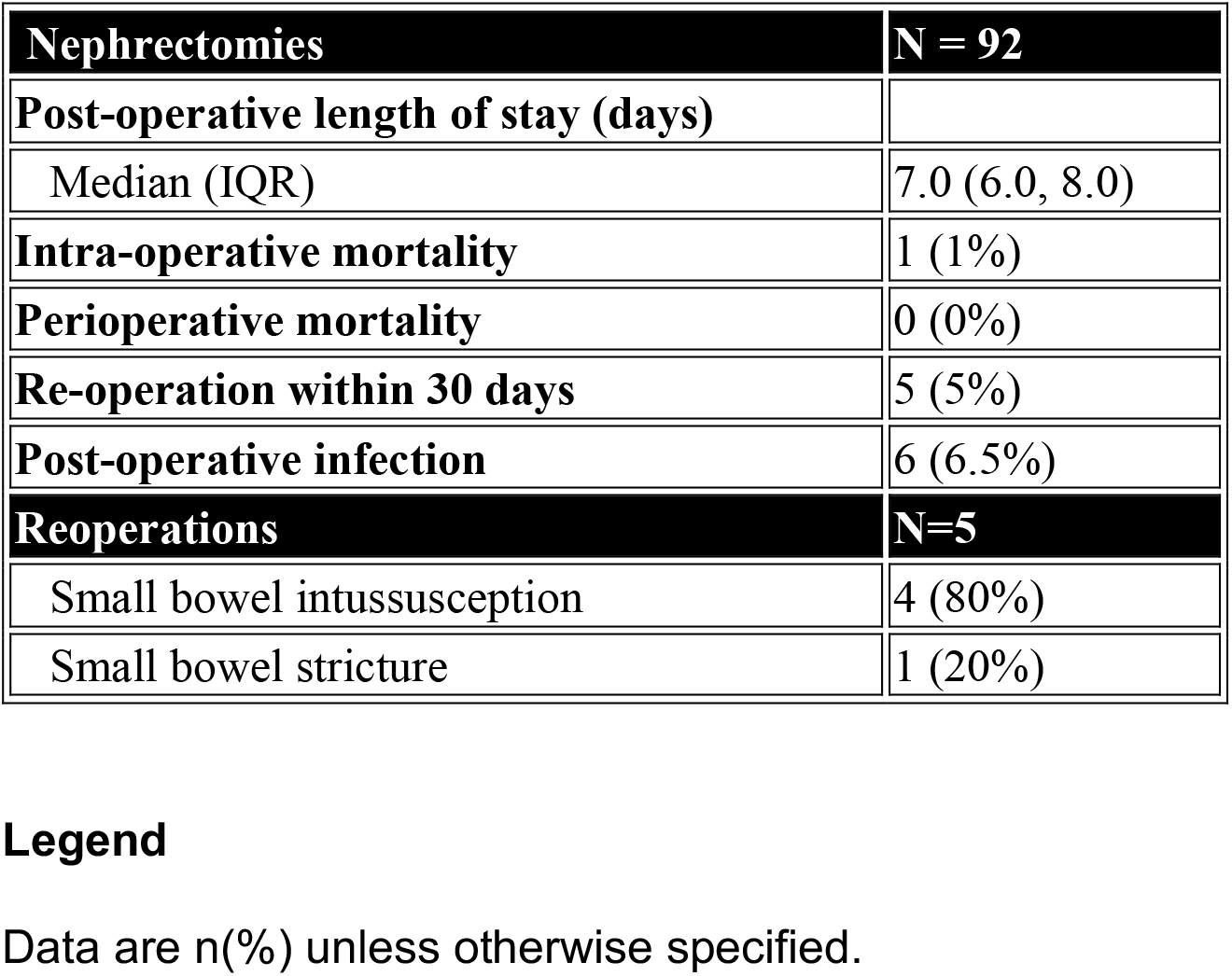
Surgical outcomes of nephrectomy.

### Survival outcomes

Median follow-up time was 21 months (IQR, 13-31). Overall survival (OS) at one and two years was 59% (95%CI, 51-69) and 48% (95%CI, 39-60), respectively. Event free survival (EFS) was 34% (95%CI, 27-43) at both one and two years (Figure 2). Among those who completed all therapy, two-year OS was 80% (95%CI, 69-93) and EFS was 69% (95%CI, 59-82). Two-year EFS ranged from 0% in those with Stage V disease to 60% (95% CI, 45-80) in those with Stage I disease (Table 4).

**Figure 2.**
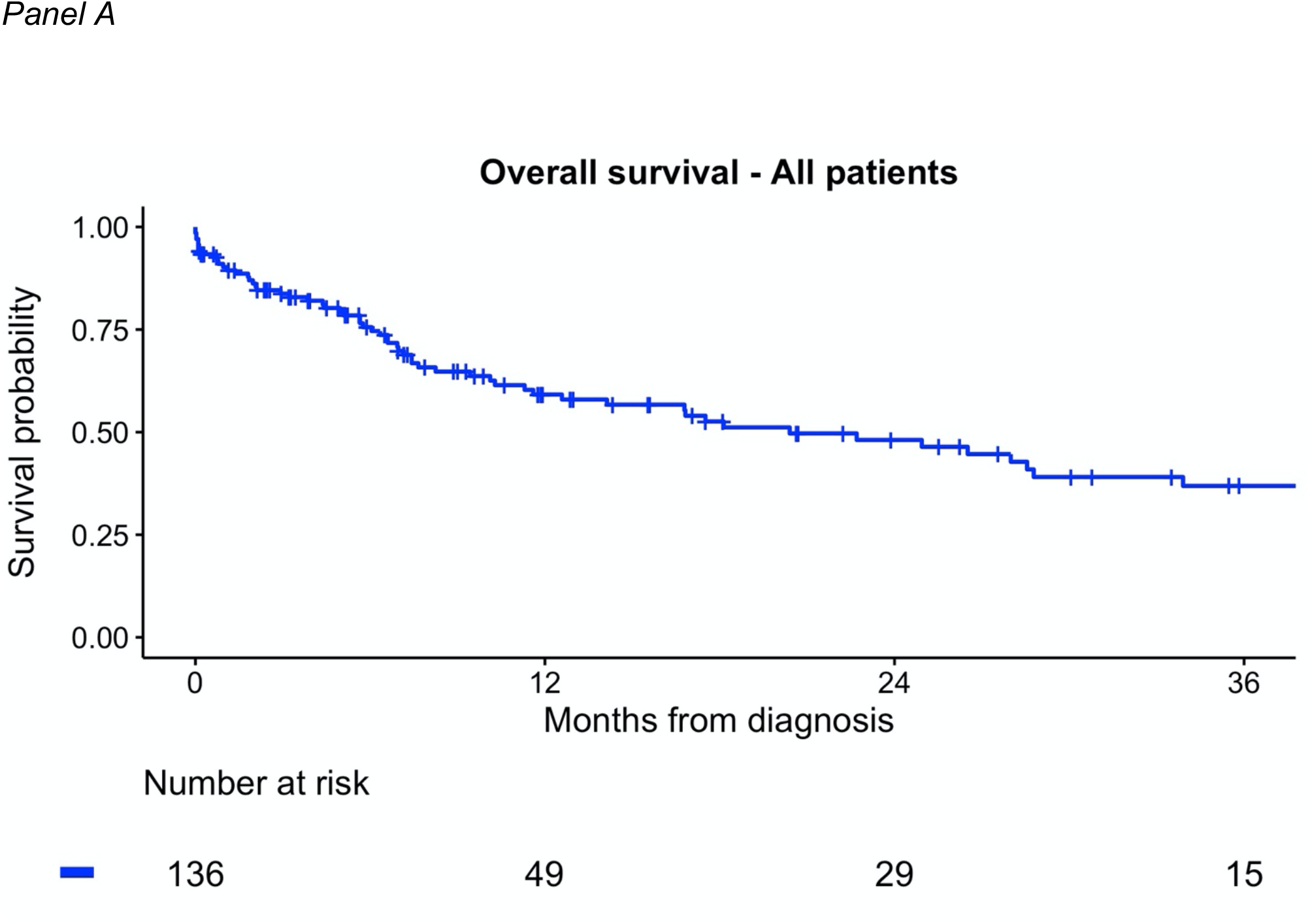
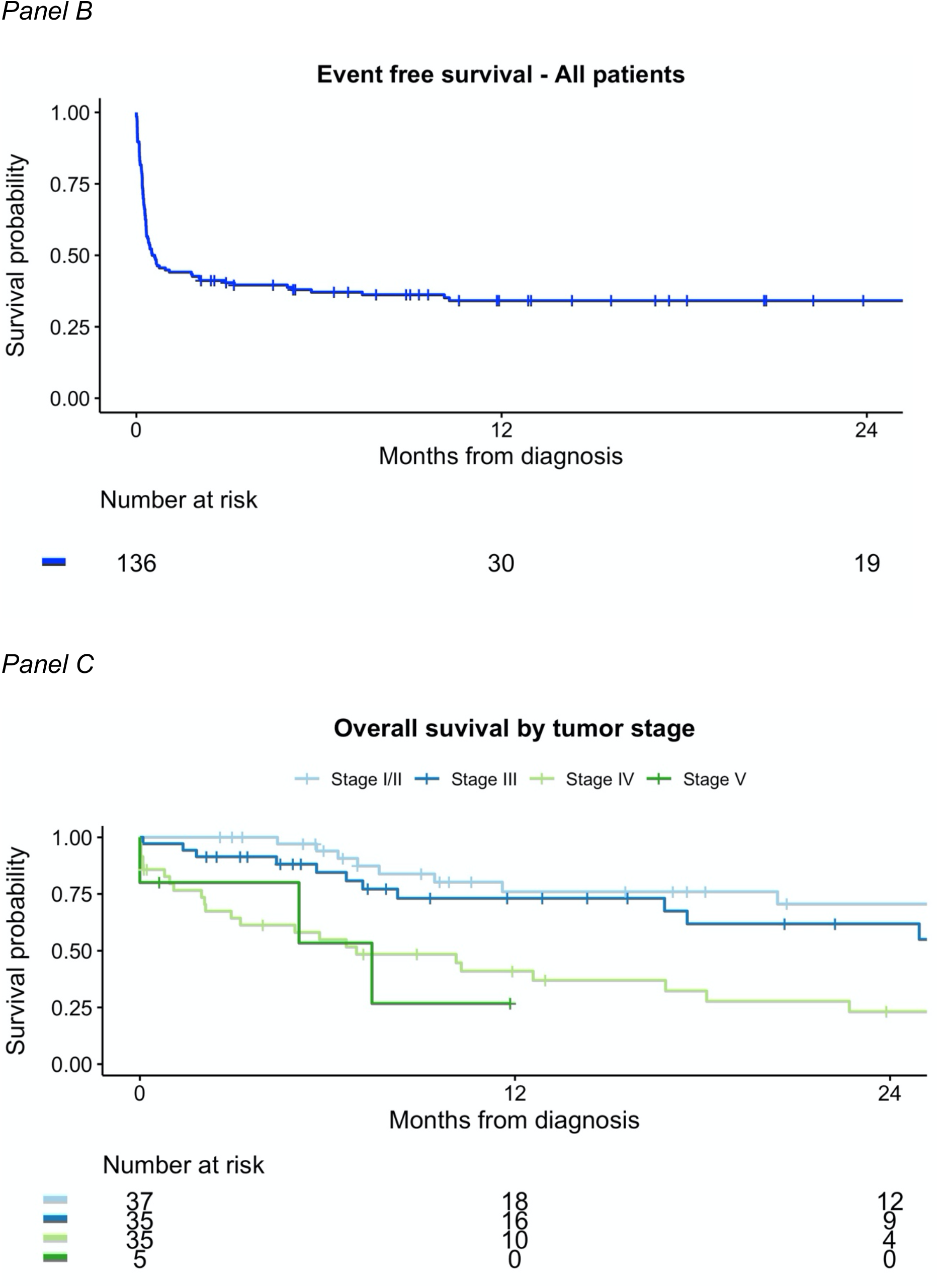
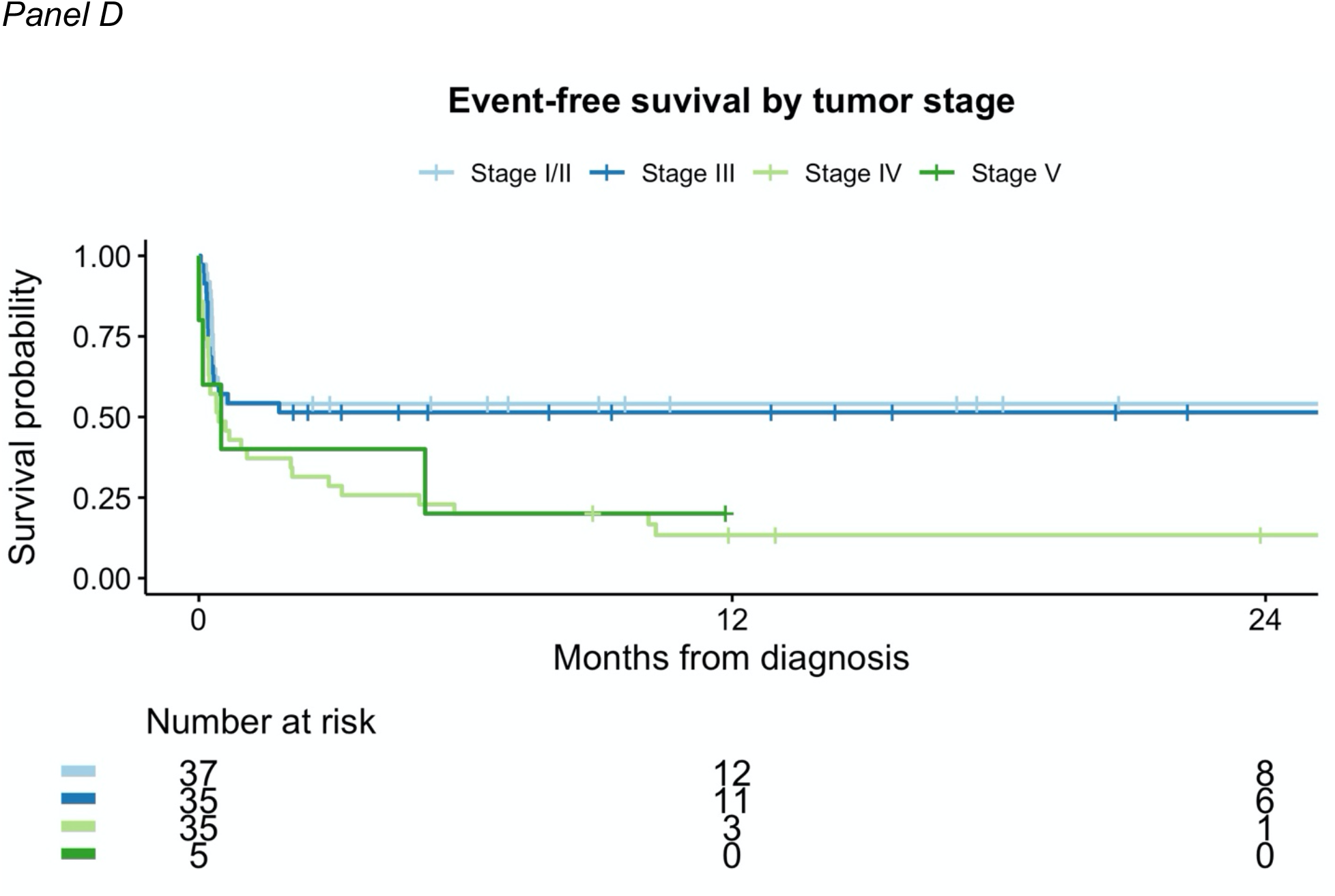
Event-free and overall survival of Wilms tumor patients in Malawi. **Legend:** Panels C & D represent 104 patients for whom staging workup was completed. Stages I & II have been combined due to low sample size among patients with Stage II disease (n=7).

**Table 4:**
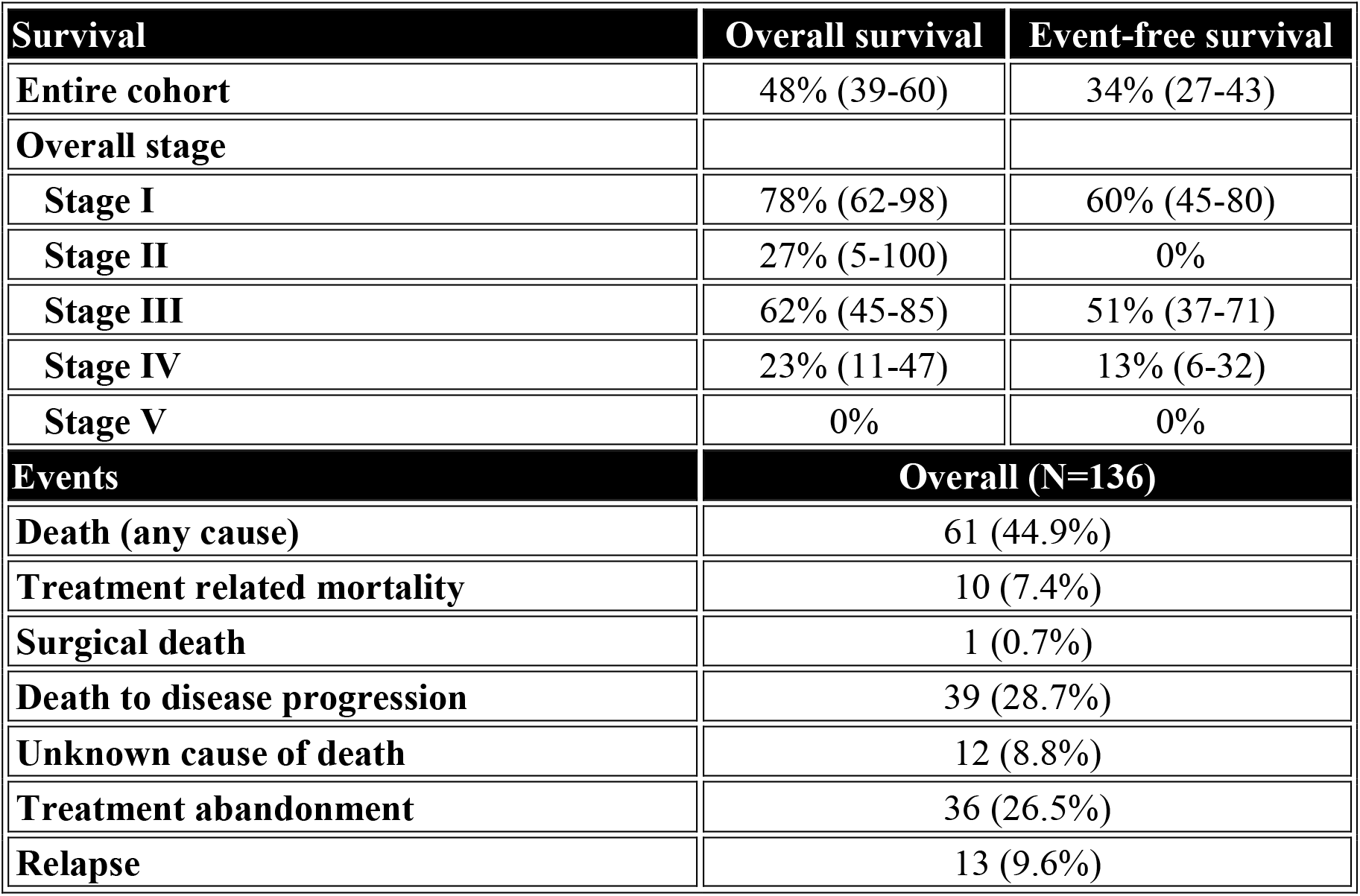
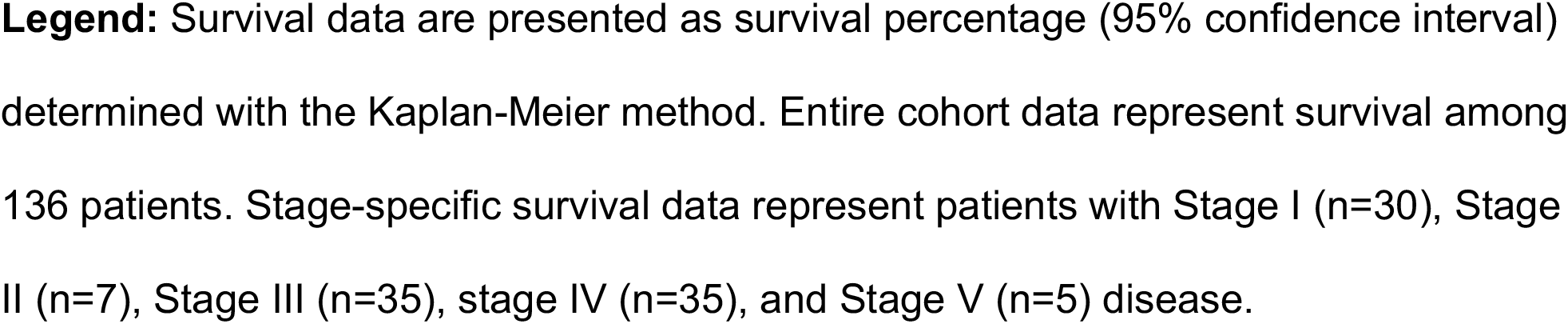
Survival and events at two years within the cohort.

| **Survival** | **Overall survival** | **Event-free survival** |
| --- | --- | --- |
| **Entire cohort** | 48% (39-60) | 34% (27-43) |
| **Overall stage** |  |  |
| Stage I | 78% (62-98) | 60% (45-80) |
| Stage II | 27% (5-100) | 0% |
| Stage III | 62% (45-85) | 51% (37-71) |
| Stage IV | 23% (11-47) | 13% (6-32) |
| Stage V | 0% | 0% |
| **Events** | **Overall (N=136)** |  |
| **Death (any cause)** | 61 (44.9%) |  |
| **Treatment related mortality** | 10 (7.4%) |  |
| **Surgical death** | 1 (0.7%) |  |
| **Death to disease progression** | 39 (28.7%) |  |
| **Unknown cause of death** | 12 (8.8%) |  |
| **Treatment abandonment** | 36 (26.5%) |  |
| **Relapse** | 13 (9.6%) |  |

**Legend:** Survival data are presented as survival percentage (95% confidence interval) determined with the Kaplan-Meier method. Entire cohort data represent survival among 136 patients. Stage-specific survival data represent patients with Stage I (n=30), Stage II (n=7), Stage III (n=35), stage IV (n=35), and Stage V (n=5) disease.

Of 24 patients presenting with lung metastases, 16 (67%) had follow-up imaging. Fourteen patients (88%) had persistent lung involvement, among whom none were alive two years from diagnosis. Both patients with resolution of lung metastases are alive. Of ten patients with liver metastases at diagnosis, a single patient survived to two years. Of patients with vena cava involvement, two of thirteen survived.

Within the cohort, 39 (28.7%) patients died from disease progression, 10 (7.4%) from treatment-related causes (e.g., sepsis), and 12 (8.8%) of unknown causes at home (Table 4). Among eight patients returning to care after abandonment, two were alive two years from diagnosis. Of all deaths, 25% occurred within fifty days of diagnosis.

Thirteen children (9.6%) had documented relapse. All were started on a salvage chemotherapeutic regimen with dose-reduced ifosfamide, carboplatin, and etoposide, and four patients had repeat surgery for debulking. The median time to relapse was 15 months from diagnosis (IQR, 13-21 months). Two relapsed patients are alive at 3.6 and 4.7 years from diagnosis, one of whom received radiotherapy.

In addition to patients with persistent metastatic disease, thirteen patients with Stage III disease received escalated therapy with Regimen M post-operatively rather than standard therapy due to absent radiotherapy. Of these, four patients (31%) were alive at 12 months, four (31%) abandoned therapy, four (31%) died, and one (8%) was lost to follow-up prior to 12 months. In comparison, of ten patients receiving standard therapy with follow-up data available to one year, nine were alive, one died, and none abandoned.

### Factors Influencing Survival

Factors significantly associated with OS on univariable analysis were severe acute malnutrition (hazard radio, HR, 1.9; 95%CI, 1.1-3.2), increasing tumor stage (HR, 1.5; 95%CI, 1.2-1.8), and vena caval tumor involvement (HR, 2.7; 95%CI, 1.4-5.4). A post-hoc multivariable analysis showed that acute malnutrition was no longer significantly associated with outcome when controlled for tumor stage at diagnosis.

## Discussion

Wilms tumor is highly curable with well-established surgical techniques, minimal-to-moderately myelosuppressive chemotherapy, and external beam radiation that can be given safely to young children. Its backbone chemotherapy is relatively inexpensive, widely available, and typically well-tolerated with minimal toxicity. Wilms tumor is thus ideally suited to be cured even in resource-limited settings. Our evaluation of outcomes of a resource-adapted Wilms tumor treatment protocol in Malawi demonstrates the potential for high rates of cure for patients with low stage disease, but also the enduring barriers to broadly realizing these cure rates in resource-limited settings.

Between 2016-2021, two-year OS among patients with Wilms tumor was 48%, consistent with contemporary outcomes in the low-resource settings of sub-Saharan Africa.^23^ During this period, regional OS ranged from 0-53% at approximately three years, averaging 44% and 27% in East and West Africa, respectively. Survival reported here is similar to that reported by Queen Elizabeth Central Hospital in Blantyre, Malawi where 2-year OS and EFS was 46%.^8^ By comparison, OS in relatively high-income South Africa averaged 76% during the same period.^23^ In the most recent SIOP and COG reports from high-income countries, OS was >80% across all risk groups, approaching 100% in low-risk disease.^4,16,24^

Treatment abandonment, occurring in 27% of patients, was the major determinant of event free survival (EFS) in the cohort. In regional studies, abandonment ranges from 3% in South Africa to as high as 35% in Sudan.^9,25,26^ A recent review of all-cancer abandonment across five African pediatric cancer centers reported that contemporary rates remain consistently above 10%.^27^

While high OS among low-stage disease reflects progress within the setting of the cancer center, EFS reflects the true burden of mortality within our cohort due to abandonment. For example, OS for stage I/II disease was 71%, but EFS was reduced to 54% due primarily to abandonment. Only two patients who abandoned therapy survived to two years after completing treatment upon return, thus abandonment is most likely fatal for patients with this outcome. Addressing abandonment must be as central to resource-adapted protocols as diagnostic protocols and risk-stratification.

Treatment abandonment is a multifaceted phenomenon caused by complex structural and socioeconomic barriers that are often beyond the control of providers and families.^27,29–32^ Financial pressures are common and include unaffordable therapy, absent transportation, and loss of parental income. Developing strategies to improve retention is a top priority among providers and organizations working in sub-Saharan Africa. For example, regional networks such as the Collaborative African Network for Childhood Cancer and Research have decreased treatment abandonment and improved outcomes for Wilms tumor through follow-up programs, robust contact networks, and direct financial support to families.^27,33^

While our center implements an active follow-up system and utilizes financial support to families, a large catchment area with travel time to the cancer center routinely >12 hours may have contributed to our high rate of abandonment, as well as incompletely addressing the financial impact of therapy for an extremely resource-limited population. Further study into implementation strategies that are generalizable to the heterogenous array of logistical and financial challenges facing cancer centers in LMICs are essential.

Another major contributor to low survival in our setting was the high proportion of patients with advanced disease at presentation. Two-thirds of staged patients had either stage III or stage IV disease, a distribution that is consistent with other African studies and is the reciprocal of the stage distribution in high-income countries where 40-60% of patients present at stages I or II.^34,35^

Addressing the negative prognostic influence of late-stage cancer presentation in low-resource settings is essential, particularly as capacity-building efforts in LMICs continue to improve outcomes among those who are able to access – and as importantly, to complete – care at these centers. At 71%, the two-year OS among patients in our cohort with stage I or stage II disease was relatively favorable in light of severe resource constraints, albeit limited by the general inability to salvage relapsed disease. By contrast, patients with stage IV disease, presenting in a quarter of patients, had an OS of 23%. Modern Wilms tumor protocols rely on high-dose chemotherapy and radiotherapy to cure late-stage disease, treatment modalities that are widely unavailable or infeasible in LMICs. Therefore, strategies to reduce late presentation in LMICs must continue to be pursued as a comprehensive approach to treating Wilms tumor in our setting.

Finally, the regional absence of radiation therapy is an important barrier to cure, evidenced by dismal outcomes among patients who required it. Among 55 patients (40.4%) for whom radiotherapy was indicated only three received it. Among patients with persistence metastases at time-of-surgery, a population where radiotherapy is critical, none survived despite escalating chemotherapy intensity. Although a larger study with sufficiently powered hypothesis tests is required to draw conclusions, increasing post-operative therapy intensity to Regimen M among patients with Stage III disease had no apparent effect on survival in our cohort. Abandonment was exclusive to patients receiving higher-intensity therapy in this sub-cohort, highlighting the potential risk of treatment intensification leading to higher rates of abandonment.

These data support SIOP’s resource-adapted recommendations that absent radiotherapy, patients with persistent metastases at time of nephrectomy should be transitioned to palliative care.^12^ In centers with access to robust pediatric surgery programs, excision of resectable metastatic lesions as implemented in the SIOP 93-01 trial may be considered in these cases, allowing for continuation with standard post-operative regimens that have been shown to result in favorable response.^36^ However, in the cases of unresectable metastatic disease at the time of nephrectomy, limited supportive care capacity in LMICs likely precludes the most myelosuppressive salvage regimens incorporated in that trial.

Successful pediatric surgical programs are essential to cure Wilms tumor. Our data show that a successful surgical oncology program is feasible in resource-limited settings, particularly among patients with low-stage disease. The overall post-operative surgical complication rate was 13%, the majority being infections treated effectively with antibiotics, thus approximating complication rates of high-resource settings.^37^

Treatment-related mortality, occurring in 16% of patients, was typical of regional experience but more common than that documented in high-resource settings.^16,26,38^ Toxicity associated with Wilms tumor is less common than that observed in treating cancers such as Burkitt lymphoma in resource-limited settings, but these high rates relative to high-resource settings underscore the continued importance of building supportive care capacity in LMIC cancer centers.

### Strengths and Limitations

This study leveraged high-quality clinical data documenting six years’ experience treating Wilms tumor in a resource-limited setting in sub-Saharan Africa to create among the largest datasets of clinical outcomes in such a setting yet published. In doing so, the study achieves stage-specific insights into the realities of treating Wilms tumor in a low-income country, and it provides support for the priorities to be pursued with next-step funding, research, and intervention. As with all retrospective studies, the study was limited by bias introduced by incomplete data collection or documentation (e.g., chest imaging). Inconsistent drug availability mandated frequent deviation from the protocol, thus creating significant heterogeneity in the approach to treating individual risk groups and limiting conclusions regarding risk-group-specific treatment approaches. However, protocol deviations due to stockouts are common in pediatric cancer care in sub-Saharan Africa, thus the protocol’s pragmatic, adaptable approach is generalizable to the broader region.^38,39^ Finally, as long-term follow-up after therapy was not maintained for many patients, rates of relapse may be underreported in this study.

## Conclusions

Survival rates of children with Wilms tumor in this low-resource setting are a stark reminder that the greatest prognostic factor for Wilms tumor survival is local treatment capacity rather than tumor biology. Capacity-building programs in sub-Saharan Africa have resulted in favorable outcomes among children with low-stage disease and those who complete therapy, but factors such as advanced disease at diagnosis, concomitant comorbidities, high abandonment rates, and lack of available radiotherapy represent the important scientific, public health, and capacity-building yet to be sufficiently addressed.

## Data Availability

All data produced in the present study are available upon reasonable request to the authors.

## Acknowledgements

We would like to thank the physicians, nurses, pharmacists, social workers, therapists, and administrators that have dedicated their professional lives to the care of children with cancer in Malawi. We thank our patients and their families for contributing their data to this manuscript.

