## Supplemental tables for "Outcomes of a resource-adapted Wilms tumor treatment protocol in Lilongwe, Malawi, 2016-2021: successes and enduring barriers to cure"

**Supplementary Table 1: Wilms tumor staging and histological grading in SIOP protocols**

| **Tumor Stage** | |
| --- | --- |
| **Stage I** | Fully resected tumor with intact renal capsule. Disease is limited to the kidney, pelvic system, and/or intrarenal vessels but does not involve the renal sinus vessels and does not extend beyond the margins of resection. |
| **Stage II** | Fully resected tumor with tumor extension beyond the renal capsule. Tumor infiltrates may extend to the renal sinus vessels, peri-nephric lymph nodes, and/or adjacent organs, but all disease must be resected with negative margins. |
| **Stage III** | Following resection, residual tumor is present but is confined to the abdomen, including lymph node involvement, locoregional spread, tumor spillage during surgery, peritoneal involvement, vena cava involvement, or pre-surgical biopsy. |
| **Stage IV** | Hematogenous spread (e.g., lung, liver) or involvement of lymph nodes outside of the abdomen or pelvis. |
| **Stage V** | Bilateral kidney involvement with or without hematogenous spread |
| **Histologic grade** | |
| **Low risk** | 100% necrosis |
| **Intermediate Risk** | Regressive, epithelial, stromal, mixed, or focal anaplastic histology |
| **High risk** | Blastemal or diffuse anaplastic histology |
| Abbreviations: SIOP, International Society of Paediatric Oncology | |

**Supplementary Table 2: The Global HOPE Wilms tumor protocol**

| **Pre-operative chemotherapy** | | | | | |
| --- | --- | --- | --- | --- | --- |
| **Pre-operative risk group** |  | | | | |
| **Unilateral, non-metastatic** | VA 4 cycles (8 weeks) | | | | |
| **Metastatic or Stage V** | VAD 4 cycles (8 weeks) | | | | |
| **Post-operative chemotherapy** | | | | | |
| **Tumor histology** | **Tumor stage** | | | | |
|  | **Stage I** | **Stage II** | **Stage III** | **Stage IV** | **Stage V** |
| **Low risk** | VA 5 cycles | VA 5 cycles | VA 5 cycles | VAD or M 5 cycles | VAD or M 5 cycles |
| **Intermediate risk** | VA 5 cycles | VA 5 cycles | VAD or M 5 cycles | VAD or M 5 cycles | VAD or M 5 cycles |
| **High risk** | VAD 5 cycles | VAD 5 cycles | VAD or M 5 cycles | VAD or M 5 cycles | VAD or M 5 cycles |
